# Experiences of a community-engaged placement model: student, educator, and community stakeholder perspectives

**DOI:** 10.64898/2026.02.05.25340232

**Authors:** Edwina Rushe, Anna McCurdy, Michelle O’Donoghue, Pauline Boland, Sarah Dillon, Donal O’Leary, Aisling Goode, Tanya McGarry, Anne Griffin, Belinda Simiyu, Ciaran Purcell

## Abstract

**Background:** The University of Limerick’s Allied Health Community Engaged Placement (CEP) model promotes interprofessional teamwork, enabling allied health students to learn with, from, and about each other through shared community placements. CEPs complement traditional clinical placements by offering socially responsive experiences, fostering community partnerships, and preparing students for diverse practice settings.

**Methodology:** This mixed method study used a qualitative dominant design, underpinned by social constructivism. Data was collected via focus groups and an online survey. Five focus groups were conducted with students (n=9), practice educators (n=6), and community partners (n=5). Reflexive thematic analysis underpinned qualitative analysis. The survey included an 8-item, 6-point Likert scale to assess perceived skill development, distributed to all three groups.

**Findings:** Six themes were generated: (i) lifting the veil of the social determinants of health, (ii) not your traditional placement, (iii) developing skills to serve communities, (iv) rolling with resistance, sense of growth, and (vi) beyond placement beyond academia. Students valued interprofessional and collaborative skills developed, though some noted clinical skills gaps. Educators observed improved communication and teamwork, while community partners highlighted students’ leadership, professionalism, and meaningful contributions to the community.

**Discussion:** Community engaged placements support development of teamwork, adaptability, and cultural responsiveness skills essential for addressing health inequities. These competencies are achieved through transformative experiences with community members, which cultivate shared learning and growth for students, practice educators, and partners. To maximise impact for all three groups guidelines are needed to support understanding, preparation, and sustainability of placements.

## Introduction

Contemporary healthcare models emphasise preventive interventions, inclusivity, and community-based care, highlighting the need for graduates who are culturally responsive, clinically skilled, and equipped to work collaboratively with communities and across professions to address the holistic determinants of health (World Health Organization [WHO], 2020, 2022). In Ireland, this is reflected in frameworks such as Sláintecare’s ‘right care, right place, right time’ and the Healthy Ireland strategy, which highlight the need for integrated services supporting the whole population with equitable geographic access (Department of Health, 2021; Health Service Executive [HSE], 2022).

Practice education is a key component of professional preparation for all health professionals, offering students authentic opportunities to apply theoretical knowledge, develop clinical reasoning, cultivate professional identity in contextualised environments, enhance interprofessional collaboration skills, and build readiness for autonomous professional practice (WHO, 2020; Higgs et al., 2019; Health and Care Professions Council [HCPC], 2023).

Interprofessional education (IPE) within placements is essential for preparing students to collaborate effectively, as outlined in the Framework for Action on Interprofessional Education and Collaborative Practice (WHO, 2010). Given the increasing complexity of healthcare needs, no single profession can adequately address the multifaceted determinants impacting upon individuals and communities, which reinforces the importance of collaborative, team-based practice across disciplines (Barr, 2015; Reeves et al., 2016).

IPE enables students to acquire the competencies necessary for effective teamwork and collaboration in healthcare settings (Oudbier et al., 2024). IPE enhances understanding of professional roles, fosters respect, and improves communication skills essential for delivery of coordinated patient-centred care. As healthcare systems strive to adopt collaborative models, IPE has become a key component of university curricula, consistent with WHO’s call to prepare practitioners capable of working interprofessionally to improve population health outcomes (WHO, 2010; Oudbier et al., 2024).

Contemporary allied health education must also prepare graduates for diverse practice contexts that extend beyond traditional clinical settings (Potvin et al., 2024). Community-engaged placements (CEPs) represent one such model, providing authentic learning opportunities in non-traditional, often underserved, community-based environments where interprofessional collaboration is pivotal (Longman et al., 2020). Robust evidence on the long-term outcomes of these placements for service users remains limited and mixed due to variables that influence interprofessional collaboration and learning (Reeves & Hean, 2013; Reeves et al., 2016).

CEPs are identified collaboratively by the university and community partners in response to local needs, where students work in settings without an established allied health professional role (Cooper & Raine, 2009). Supervision follows a blended model, combining remote guidance from profession-specific educators with on-site support from community staff (Dew et al., 2024). Interprofessional CEPs provide students with authentic opportunities to develop autonomy, adaptability, and responsiveness to the needs of the community while being embedded for the duration of placement within that very setting (Thistlethwaite, 2013).

Such experiences are underpinned by professional and international standards such as those of the World Federation of Occupational Therapists (WFOT) and the Irish Society of Chartered Physiotherapists (ISCP), which emphasise supervised practice hours to ensure public safety and the attainment of core professional competencies (WFOT, 2016; ISCP, 2021).

Interprofessionally focused community-engaged placements offer educational value by enabling students to confront the complexity, unpredictability, and social context of health and social needs, while working collaboratively across disciplines. Collaboration across disciplines is essential in this context, reflecting the complex, interdependent nature of modern healthcare delivery (Patel & Reeves, 2018). Given the pressures of modern healthcare, interprofessional collaboration is no longer optional; it is essential for coordinating patient-centred, holistic care (Patel & Reeves, 2018).

Community-based education models provide opportunities for students to apply learning in real-world contexts while developing interprofessional skills (Warren et al., 2011; Uy, 2019). CEPs promote co-designed service delivery, encouraging shared decision-making and mutual benefit for students and community partners (Furco, 1996). Evidence from practice-based IPE highlights links between such experiences and improved teamwork among future clinicians (Gilligan et al., 2014; O’Leary et al., 2019). Studies from international contexts, including Australia and the Philippines, show that community placements also enhance students’ leadership, communication, advocacy, and civic engagement (Held et al., 2019; Uy, 2019). These benefits are particularly relevant in addressing social determinants of health. Poverty and related disadvantages such as poor housing, limited education, and reduced access to healthcare contribute to health inequities and social exclusion (Social Justice Ireland, 2023; Centers for Disease Control and Prevention [CDC], 2024; Barrett et al., 2016). Growing awareness of these structural factors has reinforced the importance of collaborative, long-term partnerships between universities, communities, and services to promote sustainable, accessible and equitable health outcomes (Arcaya et al., 2016; McGowan et al., 2021).

Community engagement is reported to enhance positive health behaviours, self-efficacy, and social support particularly for socially and economically disadvantaged groups by enabling a bottom-up approach where social inclusion is at the core (O’Mara-Eves et al., 2013). Meaningful engagement fosters empowerment, resilience, and better individual and collective health (Baxter et al., 2022; Agger & Jensen, 2015). Impactful community engagement depends on understanding needs, motivation, and context, influencing long-term outcomes (O’Mara-Eves et al., 2013). Both direct beneficiaries (actively engaged individuals) and indirect beneficiaries (wider community, service providers and policymakers) benefit through improved health skills, learning, access to preventive services, and policy alignment (O’Mara-Eves et al., 2013). Allied health professionals and students have a role in building relationships, identifying unmet needs, and co-creating solutions with local partners, which aligns with national and global policies (Department of Health, 2021; WHO, 2020).

The XXX School of Allied Health began partnering with XXX AccessCampus in 2009 to deliver community-based placements. Located in South County XXX, AccessCampus is a community–university initiative developed in collaboration with the XXX Enterprise Development Partnership. It aims to bridge education and socially disadvantaged communities by offering outreach and educational resources to a wide range of groups, from children to adults. It supports learners of all ages in disadvantaged areas through education and outreach and hosts student placements, promoting interprofessional learning and community engagement. This partnership has expanded over time to involve multiple disciplines, including allied health, public health, and music therapy students. It works closely with local organisations such as DEIS schools (Delivering Equality of Opportunity in Schools), an Irish government programme that supports schools in disadvantaged areas by providing additional resources to help students facing challenges related to poverty or other difficulties, giving them a better chance to succeed in education. The partnership also involves working with groups such as older adults and migrant families and delivering co-designed educational and health promotion initiatives. This collaborative approach provides a sustainable placement model that supports interprofessional learning, helps students develop practical skills, strengthens community capacity, and addresses the social determinants of health. In the XXX School of Allied Health, these placements are interprofessional by design and delivery, aligned with the Health and Social Care Professions framework in Ireland (2021–2026), which emphasises collective identity, collaboration, and interdisciplinary working. Students from occupational therapy, speech and language therapy, and physiotherapy complete placements together, working with community members, leaders, and volunteers to deliver responsive interventions and health promotion. Interprofessional working supports the development of shared language, respect, and problem-solving, which are critical to high-quality healthcare (Wojkowski et al., 2019). These placements are especially important in regions with persistent social and health inequalities. In Ireland, 10.9% are at risk of poverty (Central Statistics Office [CSO], 2024), with one-third of XXXX residents living in disadvantaged areas (Pobal HP Index, 2023). XXXX has one of Ireland’s highest concentrations of poverty (HSE, 2024; Limerick City and County Council, 2023). These inequalities affect health outcomes, access, and participation, highlighting the need for multidisciplinary health professionals to engage with this underserved community.

Research has shown that non-traditional practice placements increase autonomy, confidence, and professional identity (Bossers et al., 1997; Carey & Mechefske, 2016), but most literature focuses primarily on the student experience only. This emphasis means that perspectives from other key stakeholders, such as practice education staff and community members, are often underrepresented. Some acknowledge the importance of context and exposure to social issues (Dancza et al., 2013), but gaps remain in understanding how interprofessional, community-based placements function as shared learning among students, discipline supervisors, and community partners. Despite growing placement provision in Ireland, limited research explores the interconnected experiences from an interprofessional perspective.

This study explores the experiences of allied health students from occupational therapy, physiotherapy, and human nutrition and dietetics, alongside their discipline-specific educators and community partners. It seeks to understand the value of these placements and identify opportunities for enhancing practice education within non-traditional, community-based settings.

## Method

### Study Design

This mixed methods study was guided by a social constructivist approach, with qualitative dominance (Schoonenboom & Johnson, 2017). Meaning was recognised as co-constructed through interaction and dialogue (Creswell & Poth, 2018). Reporting followed COREQ guidelines (Tong et al., 2007; Booth et al., 2014; Naeem et al., 2023). Qualitative data were drawn from five focus groups conducted either online or face-to-face to facilitate interactive discussion (Carey & Asbury, 2012). Participants included students, practice educators, and community partners. Students completed a face-to-face session at the end of placement (facilitated by CP, who was independent of assessment), while educators and community partners participated online for convenience. Discussions explored experiences and reflections on community-engaged placements .A post-placement survey was distributed to all participants. It comprised eight Likert-scale items (1 = strongly disagree to 6 = strongly agree) assessing perceptions of skill development in practice skills, leadership, professionalism, teamwork, collaboration, independence, communication, and quality care. These competencies reflect cross-disciplinary professional standards in Irish allied health education and align with CORU standards and WHO interprofessional collaboration frameworks (World Health Organization, 2010).

### Recruitment

Purposive sampling targeted students from occupational therapy, physiotherapy, and human nutrition and dietetics who had completed CEPs at the XXXX between (2021–2024), their supervising practice educators, and community partners. Invitations were distributed via email, and participants who met inclusion criteria were enrolled. A semi-structured topic guide was developed and piloted. Each focus group was co-facilitated by two researchers one leading the discussion and one moderating and conducting member checking.

### Ethical Considerations

Ethical approval was granted by the Faculty of Education and Health Sciences Research Ethics Committee, University of XXX (Ref: EHSREC 2023_03_09_EHS).

### Data Management

Five focus groups (48 minutes on average) were audio-recorded, transcribed verbatim, and anonymised. Two researchers independently reviewed transcripts. Survey data were collected and stored securely using Qualtrics and password-protected OneDrive in line with GDPR protocols.

### Focus Group Thematic Analysis

Data were analysed using Braun and Clarke’s (2022) reflexive thematic analysis to identify and interpret patterns of meaning. NVivo supported data management. Coding was inductive, following their six-phase framework, with iterative team reviews carried out to refine and consolidate themes.

**Table 1.**
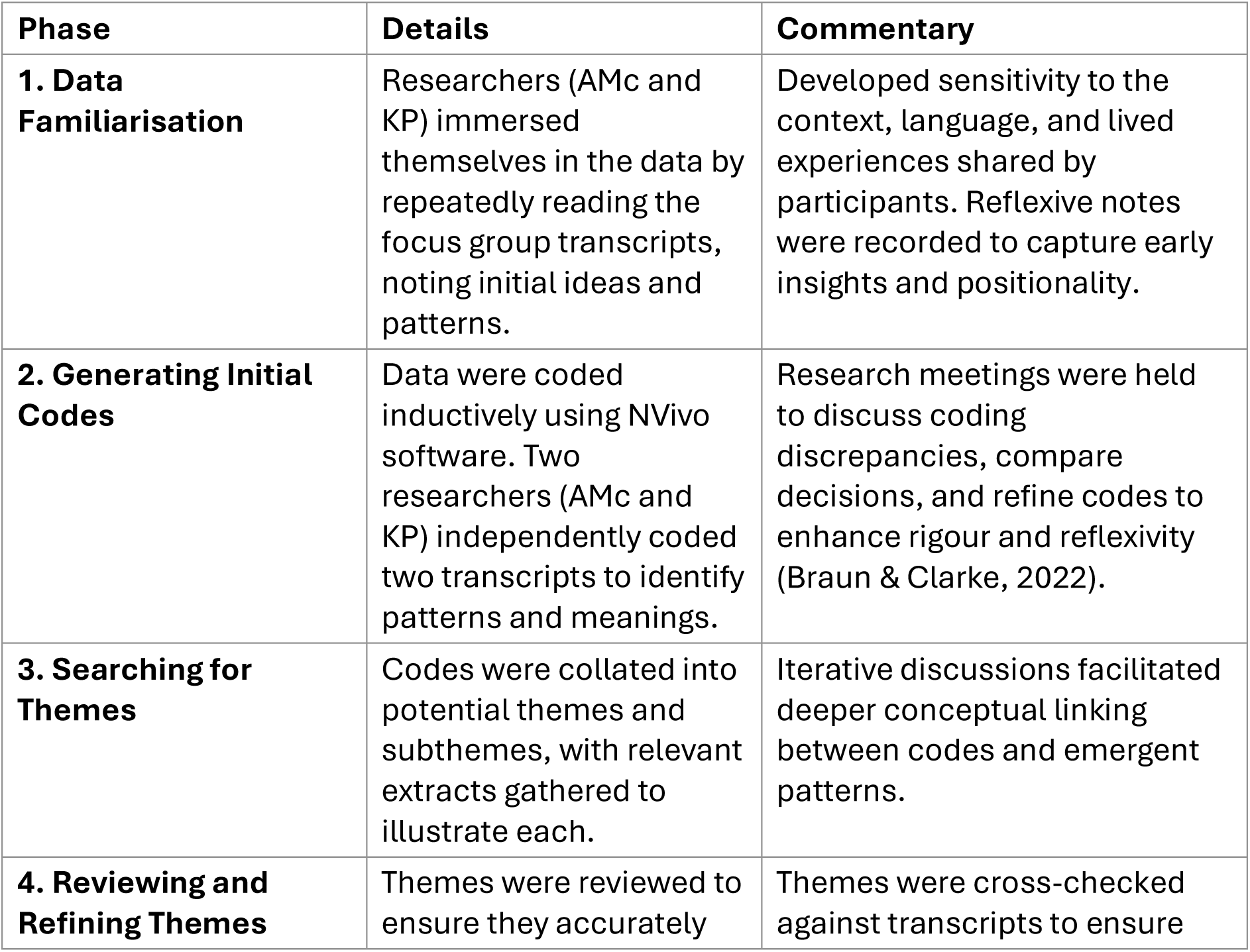
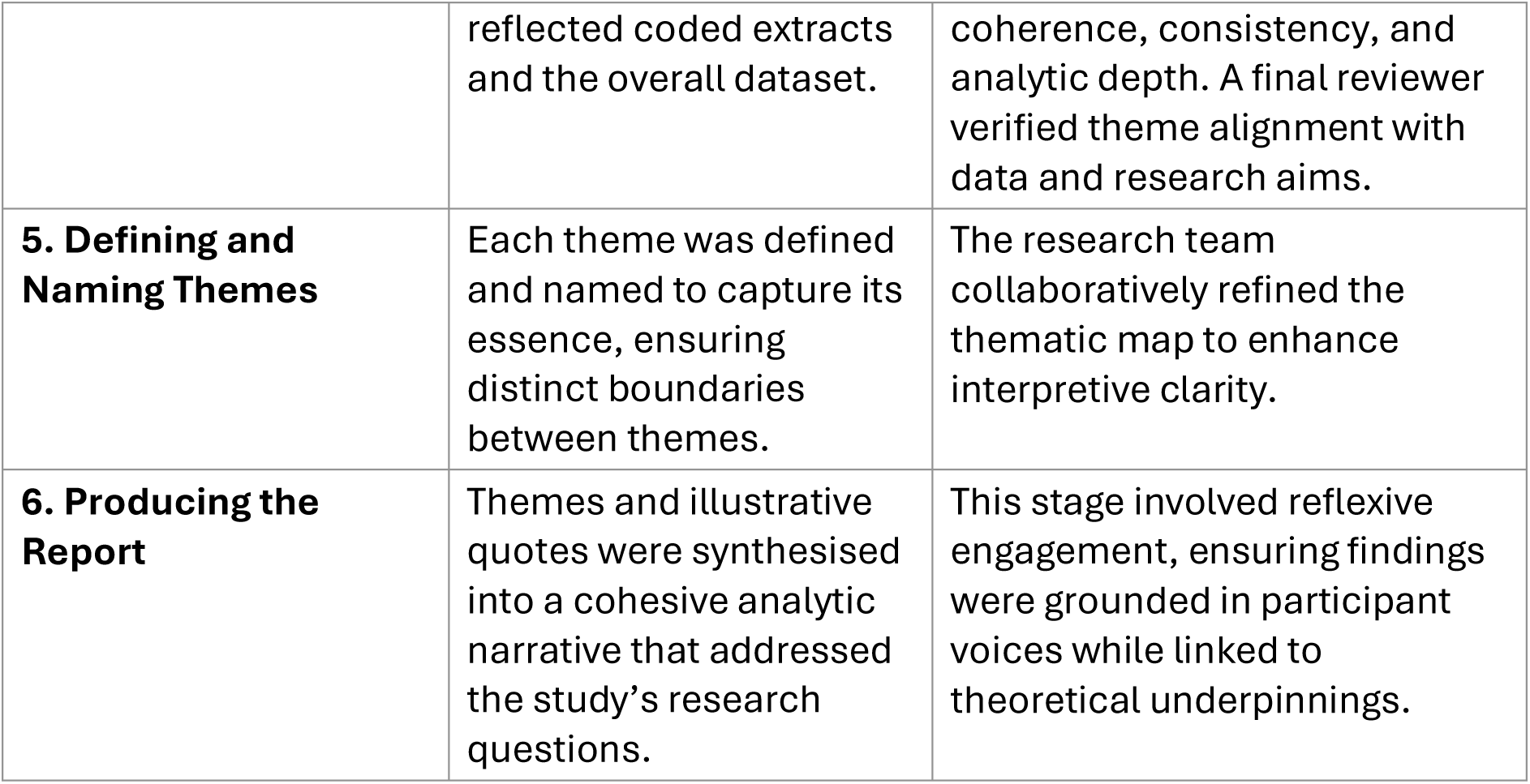
Reflexive Thematic Analysis Steps (Braun & Clarke, 2022)

### Survey Analysis

Descriptive statistics (frequencies and percentages) were calculated to identify trends in perceived competency development and to compare areas of consensus or variation across groups.

### Positionality and Trustworthiness

The research team brought experience across allied health education, community engagement, and clinical practice. Positionality was acknowledged as influencing interpretation (Berger, 2015). Reflexivity was maintained through regular meetings and supervision (Braun & Clarke, 2023). Trustworthiness adhered to Lincoln and Guba’s (1985) criteria for credibility, transferability, dependability, and confirmability.

**Table 2.** Trustworthiness Criteria (Lincoln & Guba, 1985)

| Criterion | Description |
| --- | --- |
| **Credibility** | Achieved through reflexivity engagement with data, and triangulation of researcher perspectives (AMc and KP). Regular debriefing sessions supported critical reflection and minimised bias during data analysis. |
| **Dependability** | Maintained through an audit trail detailing all stages of data management, including focus group facilitation, transcription, coding, and theme development, ensuring procedural transparency. |
| **Confirmability** | Established via reflexive journaling and peer review, which enhanced awareness of researcher influence and supported interpretive transparency. |
| **Transferability** | Strengthened by detailed contextual descriptions of settings, participants, and methods, allowing readers to evaluate the applicability of findings to other contexts. |

## Findings

This study involved three key stakeholder groups: Practice Educators (PE), Students (S), and Community Members (CM). These abbreviations are used throughout the quotes, along with focus group numbers (e.g., FG1, FG2) to indicate the source of each response.

**Table 3.** Participant Demographics.

| **Stakeholder Group** | **Number of Participants** | **Professions / Roles** | **Years of Supervision / Experience** | **Focus Group Involvement** | **Other Details** |
| --- | --- | --- | --- | --- | --- |
| **Students (S)** | 9 | Occupational Therapy (3), Dietetics (4), Physiotherapy (2) | First placement experience | One focus group (9 students) | Allied health students in Community-Engaged Placements (CEPs). |
| **Practice Educators (PE)** | 6 | OT (4), Dietetics (1), Physio (1) | Median = 9 years | Three focus groups (two per group; one pilot) | Focused on professional supervision. |
| **Community Members (CM)** | 5 | Community staff, liaison staff, volunteers | Median = 10 placements supported | One focus group (5 community members) | Represented a range of community partners supporting placements. |

**Table 4.**
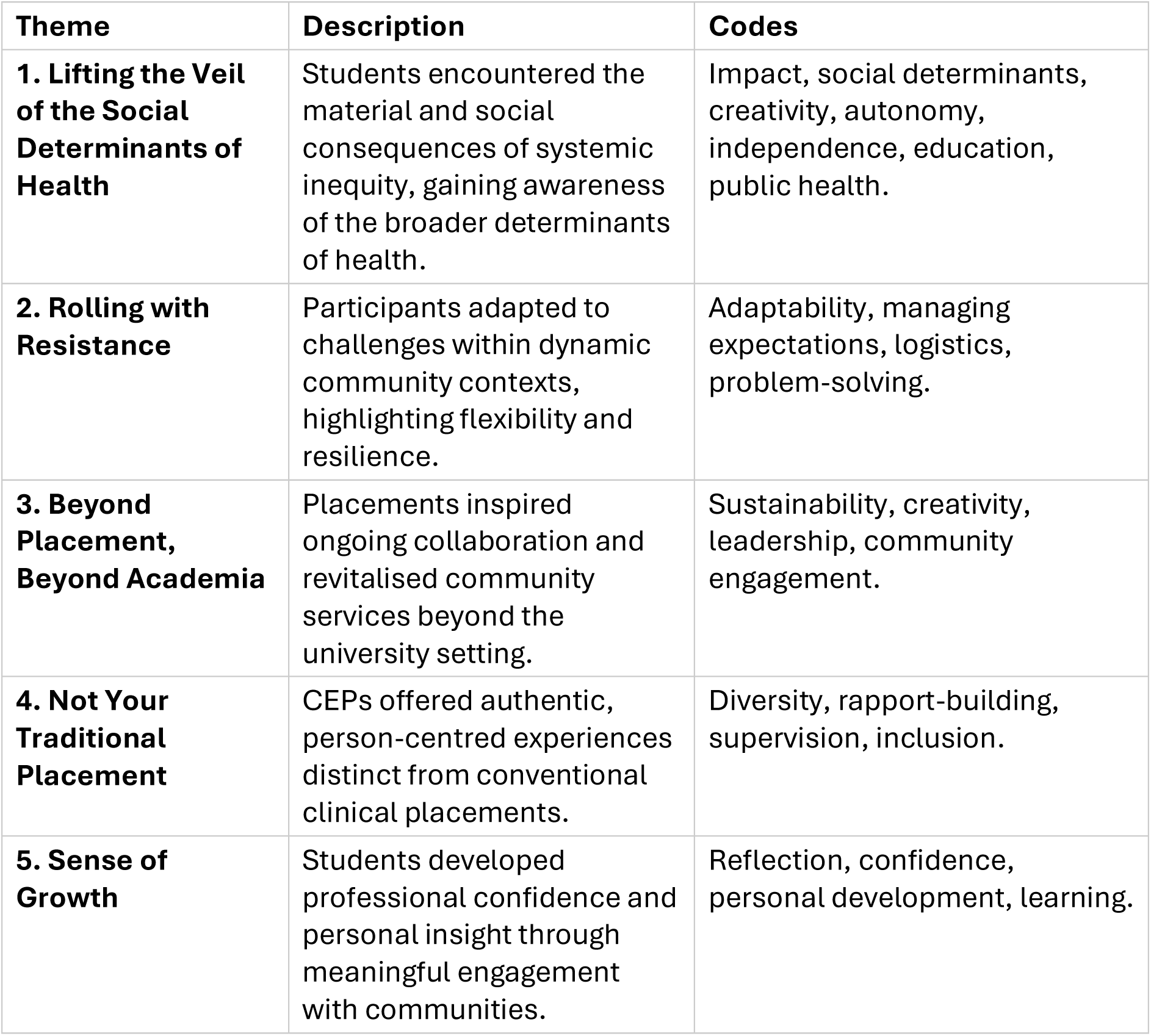
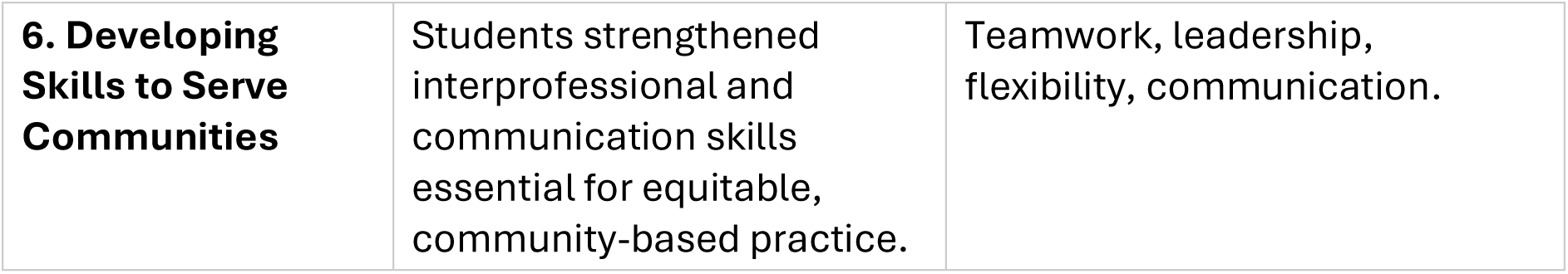
Theme Generation.

### Lifting the Veil of the Social Determinants of Health

CEPs allow students direct opportunities to meet, understand, and support people experiencing area-based deprivation, offering a first-hand view of how poverty impacts upon health. While students entered placement with theoretical knowledge of social determinants of health, the realities challenged their views and assumptions.

One educator shared how students were surprised by infrastructural constraints “Students realised there was no supermarket in [this area]… they just took it for granted that everyone drives,” and failed to anticipate that “a lot of the men in the men’s shed didn’t drive… some people rely on public transport.” (PE05, FG5). These challenges showed how interventions without awareness of the context may miss their target needs. A community member added that community educational factors also shape how health information is received: “Because the Traveller women… maybe can’t read… they wouldn’t understand nutrition and what’s healthy.” (CM03, FG2). Students began to see how barriers were embedded in economic conditions. They recognised these weren’t isolated but part of broader inequality. “We had to account for a lot of people’s social circumstances… some of their siblings are even in prison… so many things you wouldn’t usually consider.” (S06, FG4). One student reflected: “You realise very quickly that not everyone has the same choices. It’s not just about telling people what to eat, it’s what they can afford, what’s near them, and what they understand.” (S07, FG4). Educators observed how these realisations shifted thinking: “One student said, ‘we talk about health equity in lectures, but here, I saw what that meant for real.’” (PE01, FG1).

Community partners saw this awareness build advocacy “Sometimes people don’t speak up because they feel they’re not entitled. The students helped give a voice to those people.” (CM02, FG2)

### Not Your Traditional Placement

CEPs create a relational learning environment grounded in the community’s experience, making them a distinct type of placements. One community member explained the strength of CEPs lies in connections “It’s not about bringing the academic… but engaging with the person on the ground… and the students did that… they brought their own stories and that really… it’s trust.” (CM03, FG2). This allowed for deeper relationships to develop. For students, it meant learning to meet people where they are emotionally and culturally.

Educators and community members reflected on how CEPs opened access to groups often underserved.

A practice educator shared: “Building relationships with… community groups we wouldn’t have reached otherwise” allowed students to “make a difference… to maybe groups that wouldn’t have access.” (PE01, FG1). The way placements are set up encourages students to adapt. “There wasn’t a clear schedule,” one student said, “but it made us think on our feet.” (S04, FG4).

This flexibility isn’t usually expected of students. One educator said, “They learned to be human first, practitioner second.” (PE03, FG3). CEPs encouraged a range of cross-professional collaboration. “For two sessions out of five, we had the physios come in too. That’s how we collaborated.” (S08, FG4)

### Developing Skills to Serve Communities

Students in CEPs work with allied health professionals and diverse community groups collaboratively. One community member stated, “They must do multidisciplinary work. It might not be with a health discipline… they need to learn that.” (CM03, FG2).These interactions move students beyond their own disciplinary comfort, introducing them to youth workers, sports organisations, and community leaders, each with unique values and styles. “They worked with youth workers, teachers… even the GAA club. That’s how it is in the real world.” (CM01, FG2).

Educators reflected on shared learning: “We work as a team… then the students, with the education team and community, come up with solutions.” (PE03, FG3). Co-designing interventions gives students ownership and autonomy. One student said, “It felt like the first time we were treated like professionals.” (S03, FG4). An educator stated “They had to take initiative, use judgment, and shape the sessions. That’s a real skill.” (PE05, FG5). This dynamic model builds confidence “After doing this placement, you would feel so much more confident.” (S08, FG4). Community staff noted development of skills in adaptability: “They didn’t wait to be told. They asked, listened, and acted.” (CM02, FG2).

### Rolling with Resistance

Students, educators, and communities learned to adapt to resistance. One educator described managing changing plans “Just managing expectations… of having a plan and it suddenly changing… but that’s also the exciting bit.” (PE03, FG3). Students overcame barriers to connection too. One shared, “He asked if I was a Traveller. When I said no, he walked off. Later, I used boxing as a way in.” (S04, FG4).

Community members noted Travellers often aren’t made to feel welcome. “Sometimes you’ll find they’re not always made to feel welcome.” (CM03, FG2). Students realised trust takes time. “At first, I thought, ‘Why won’t they join in?’ But then I realised they’ve had people come and go. We had to prove we cared.” (S02, FG4). Educators noted students learned about the importance of authentic trust building “Students were surprised by how hard it was but learned to do it authentically.” (PE01, FG1). Resistance was rooted in historical prejudice. “Resistance isn’t rejection. It’s about history, dignity. Once they saw students weren’t judging, things changed.” (CM03, FG2)

### Sense of Growth for All Stakeholders

Growth extended beyond students. Community members felt energy working with students and educators gained confidence in new environments. One said, “That’s a strong benefit… there’s a lot of learning in it for us too.” (PE03, FG3). Community members saw new initiatives starting “They introduced gardening, which has become a big thing. Some of our kids go to the allotment now.” (CM04, FG2) Students also reported gaining skills for their careers “I can run a group of 30, that’s a skill you wouldn’t usually develop.” (S08, FG4). “You’re not just learning how to ‘do the job.’ You’re learning who you are in that job.” (S06, FG4) “It reminded me why I got into this field. Their energy is contagious.” (PE03, FG3)

### Beyond Placement, Beyond Academia

CEPs go beyond delivering services they help students build skills for both clinical and community work. As one community member said, “The children were showing them how to garden… that was fed back into the city council.” (CM02, FG2). Students wanted to make an impact, not just pass a placement “You want to leave something behind those matters.” (S05, FG4). Educators saw this too, “They didn’t just complete a placement. They planted ideas that are still growing.” (PE01, FG1). Another community member agreed, they helped us shape something lasting.” (CM01, FG2)

### Post-placement survey

The post-placement survey highlighted differing perspectives among the three stakeholder groups regarding their perceived development of professional and interprofessional competencies during the CEP. These results demonstrate differences in how students, practice educators, and community members viewed their growth in professional and interprofessional skills throughout the placement. presents the results of the survey collecting perceptions of students (n=9), educators, (n=6) and community members (n=5) regarding their agreement with the development of key competencies and skills during community engaged placements. Note %s have been rounded to the nearest whole number for readability.

**Table 5.**
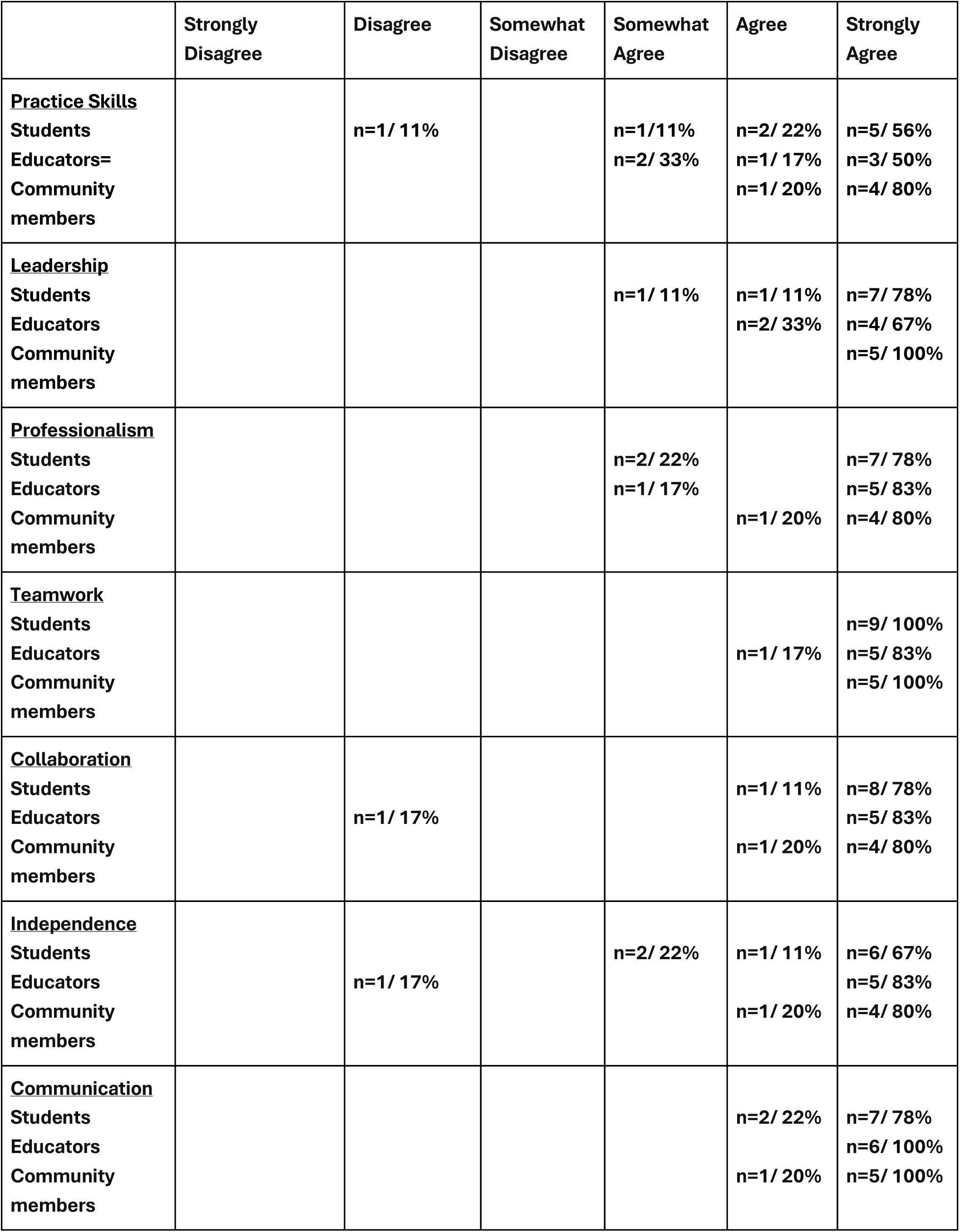
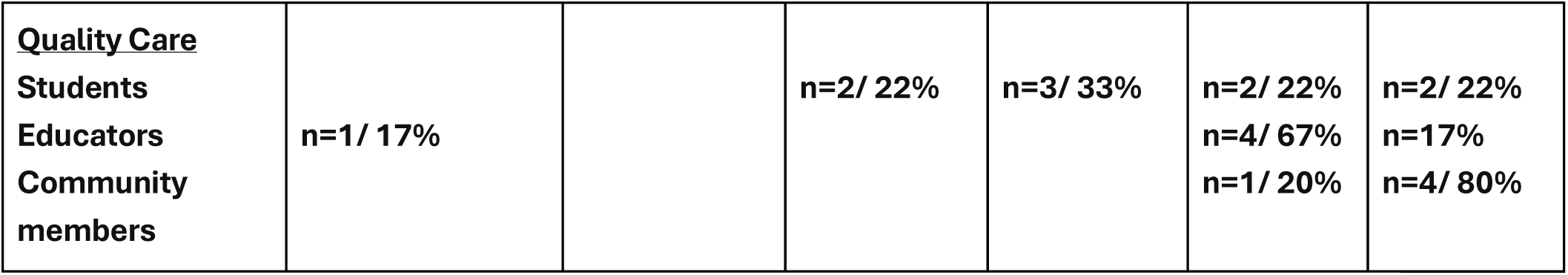
Development of Professional and Interprofessional Skills.

|  | Strongly Disagree | Disagree | Somewhat Disagree | Somewhat Agree | Agree | Strongly Agree |
| --- | --- | --- | --- | --- | --- | --- |
| **<u>Practice Skills</u>**<br>Students<br>Educators=<br>Community members |  | n=1/ 11% |  | n=1/11%<br>n=2/ 33% | n=2/ 22%<br>n=1/ 17%<br>n=1/ 20% | n=5/ 56%<br>n=3/ 50%<br>n=4/ 80% |
| **<u>Leadership</u>**<br>Students<br>Educators<br>Community members |  |  |  | n=1/ 11% | n=1/ 11%<br>n=2/ 33% | n=7/ 78%<br>n=4/ 67%<br>n=5/ 100% |
| **<u>Professionalism</u>**<br>Students<br>Educators<br>Community members |  |  |  | n=2/ 22%<br>n=1/ 17% | n=1/ 20% | n=7/ 78%<br>n=5/ 83%<br>n=4/ 80% |
| **<u>Teamwork</u>**<br>Students<br>Educators<br>Community members |  |  |  |  | n=1/ 17% | n=9/ 100%<br>n=5/ 83%<br>n=5/ 100% |
| **<u>Collaboration</u>**<br>Students<br>Educators<br>Community members |  | n=1/ 17% |  |  | n=1/ 11%<br>n=1/ 20% | n=8/ 78%<br>n=5/ 83%<br>n=4/ 80% |
| **<u>Independence</u>**<br>Students<br>Educators<br>Community members |  | n=1/ 17% |  | n=2/ 22% | n=1/ 11%<br>n=1/ 20% | n=6/ 67%<br>n=5/ 83%<br>n=4/ 80% |
| **<u>Communication</u>**<br>Students<br>Educators<br>Community members |  |  |  |  | n=2/ 22%<br>n=1/ 20% | n=7/ 78%<br>n=6/ 100%<br>n=5/ 100% |
| **Quality Care** |  |  |  |  |  |  |
| **Students** |  |  | n=2/ 22% | n=3/ 33% | n=2/ 22% | n=2/ 22% |
| **Educators** | n=1/ 17% |  |  |  | n=4/ 67% | n=17% |
| **Community members** |  |  |  |  | n=1/ 20% | n=4/ 80% |

Community members offered the most positive assessments of perceived student performance. They praised the development of leadership, communication, professionalism, and teamwork attributes that are observable in community-based, collaborative settings. All five-community staff either agreed or strongly agreed that students demonstrated strong leadership, teamwork, and communication skills. In areas such as independence, collaboration, and quality care, community staff reported predominantly positive impressions indicating that these placements contributed meaningfully not only to student learning but also to service delivery and for the community.

Students provided varied responses. While all students agreed or strongly agreed that they had developed teamwork skills, one student reported that they did not develop practice skills, and three students 39% felt they had not fully developed competencies in quality care.

Practice educators recognised student growth in areas such as communication and teamwork, but the survey raised the need for further development in practice specific competencies. One third (n=2) of educators indicated that practice skills remained underdeveloped (voting somewhat agree) by the end of the placement and one educator (17%) expressed disagreement regarding the development in collaboration, independence, and quality care.

## Discussion

### Responding to contemporary priorities

Contemporary healthcare emphasises prevention, equity, and community-based practice, calling for socially responsive and place-based learning models within allied health education. CEPs offer students opportunities to engage directly with communities, address social determinants of health, and understand experiences of marginalisation and inequality (O’Leary et al., 2020; Wilcock & Hocking, 2015). This study found that CEPs provide impactful learning experiences for students, educators, and community members, fostering awareness of the social contexts shaping health and encouraging advocacy for equity of access. These placements help to bridge health and education, positioning universities as active contributors to community wellbeing, and societal change. By placing students in non-traditional environments, CEPs bring theory to life and align with the need for socially accountable healthcare practitioners of the future (World Health Organization, 2013). In addition this model reflects the global agenda set out in the United Nations Sustainable Development Goals, particularly Goal 3: Good Health and Wellbeing, Goal 4: Quality

Education, and Goal 10: Reduced Inequalities, by promoting equitable access to healthcare, inclusive learning, and social justice through education (United Nations, 2015).

### Competency development

While the overall value of CEPs was evident, perspectives on specific competency development varied across stakeholder groups. Community members consistently praised students’ strengths in leadership, communication, and teamwork. However, both students and educators identified areas requiring further development, particularly in collaboration, independence, and quality care. These differences reflect challenges reported in interprofessional placement literature, where uncertainty about roles can shape perceptions of competence (O’Leary et al., 2019). Although community members observed students acting independently and contributing meaningfully, students and educators may have viewed these experiences through a traditional clinical lens. Some students also found it difficult to articulate non-traditional skills such as group facilitation, reflecting ongoing ambiguity surrounding interprofessional roles. This aligns with findings from role-emerging placement literature, which note that students must actively construct their professional identities in unstructured and dynamic environments (Dancza et al., 2013; 2019). These insights highlight the need for structured support to help stakeholders navigate the complexity of CEPs.

### Embracing complexity

CEPs expose students to real-world challenges such as housing insecurity, discrimination, and service gaps often invisible in traditional placement settings where service pathways are well established. Despite these challenges, students reported growth in interprofessional teamwork and adaptability, supported by the development of resilience, communication, and cultural responsiveness. These competencies are vital in community-focused practice, fostering openness, trust, respect, and empathetic dialogue (Flood et al., 2019). Consistent with the theme of *Rolling with Resistance*, students learned that trust-building requires time and authenticity, recognising resistance as a response to historical marginalisation rather than disengagement. Community members also emphasised the relational nature of intervention, which aligns with literature identifying reflective practice and interpersonal skills as foundational to care (Kyte et al., 2018; WHO, 2013).

### Structuring support and collaboration

The variability in reported skill development underscores the need for structured support, particularly for early-stage learners in non-traditional contexts. Group supervision and interprofessional learning activities allow students to learn with, from, and about each other, enhancing teamwork and communication (WHO, 2010). Embedding structured collaboration between universities and communities supports students in navigating uncertainty and achieving meaningful learning (Allan et al., 2011). Reflective opportunities throughout placement preparation, implementation, and review further enhance skill development and outcomes. The Interprofessional Practice-Based Education Profile (O’Leary et al., 2023) promotes team-based reflection and periodic review, offering a structured framework for sustainable learning environments. Building strong partnerships between universities and clinical educators is also key to ensuring mutual engagement and shared understanding of placement objectives (O’Leary et al., 2020).

### Impact

CEPs deliver benefits that extend beyond student learning. They can strengthen community partnerships and contribute to small but meaningful initiatives that continue beyond the placement period. Previous studies have shown that collaboration between students and communities can result in co-developed resources and services that persist after placements conclude (Strasser et al., 2009; Campbell, Baikie & Ranger, 2014). Educators also benefit through more facilitative roles, while community members often report a sense of shared achievement and renewed engagement. This triadic model of student, educator, and community collaboration reflects values of mutual benefit and meaningful engagement. As Warren et al. (2011) note, such placements not only support interprofessional skill development but also foster long-term partnerships that enhance local capacity.

### Limitations

This study involved a small sample within one community, limiting the transferability of findings beyond this context. Its cross-sectional design also captures perceptions at a single time point and does not reflect longer-term outcomes. Future longitudinal research should explore how CEPs influence graduates’ interprofessional practice, community partnerships, and service delivery over time.

## Conclusion

This study highlights the important role of community-engaged placements in allied health student education. These placements give students the chance to develop teamwork, communication, empathy, and adaptability while working directly with communities. They help students understand real-world health challenges and prepare them to work in diverse and complex settings. By planning placements carefully, providing strong supervision, and encouraging reflection, universities can make CEPs more effective and meaningful. Future work should focus on understanding the long-term benefits of these placements and finding ways to make them sustainable across different settings.

## Focus Group Information Students

### The topic

During your degree at UL you completed/ are completing a community engaged placement (CEP) This study is investigating the learning opportunities during this placement in comparison with other clinical education experiences.

### Ground Rules for the Group

There are some ground rules for our group, but not too many.

Firstly, <u>everyone’s experience is valid and valued</u>. We ask that all participants respect different perspectives and viewpoints and contribute to an open sharing of ideas. We would invite you all to share the full extent of your experiences including positives, negatives and areas to work on in the future as this research will inform future practice.

Secondly <u>this focus group is confidential</u> and whilst the session will be recorded participants will be deidentified and anonymised when the results are presented. We ask that all information shared is done so in full confidentiality and remains within the room.

This focus group is about having a conversation with your peers about your experiences. There are some guiding questions and prompts throughout the session but for the most part we will be guided by you and your discussions. Every now and again an interviewer may interrupt to ask a question or clarify a point.

## Operational Definitions

For this focus group, community engaged placements refer to placements where students work collaboratively with clients within their local communities. Placement settings are diverse and may include local schools, community facilities or volunteer organisations. When completing a community engaged placement students work with community members in areas to promote and enhance participation, engagement and integration within the community as well as delivering preventative healthcare and health promotion services.

Your placement was conducted over the past two years, and part or all of the placement may have occurred in Access Campus UL, in local schools or in various services based in the locality. Your placement will have involved working with different community groups and may have involved working with students from other allied health professions and other professionals. Please take a minute or two to recall that experience.

## Focus Group Information Practice Education

### The topic

Over the past number of years, you will have supervised students from UL as they completed a community engaged placement (CEP) This study is investigating the learning opportunities experienced during this placement in comparison with other clinical education experiences.

### Ground Rules for the Group

There are some ground rules for our group, but not too many. Firstly, <u>everyone’s experience</u> <u>is valid and valued</u>. We ask that all participants respect different perspectives and viewpoints and contribute to an open sharing of ideas. We would invite you all to share the full extent of your experiences including positives, negatives and areas to work on in the future as this research will inform future practice.

Secondly <u>this focus group is confidential</u> and whilst the session will be recorded participants will be deidentified and anonymised when the results are presented. We ask that all information shared is done so in full confidentiality and remains within the room.

This focus group is about having a conversation with your peers about your experiences. There are some guiding questions and prompts throughout the session but for the most part we will be guided by you and your discussions. Every now and again myself or the moderator (Sarah) will interrupt to ask a question or clarify a point.

## Operational Definitions

For this focus group, community engaged placements refer to placements where students work collaboratively with clients within their local communities. Placement settings are diverse and may include local schools, community facilities or volunteer organisations. When completing a community engaged placement students work with community members in areas to promote and enhance participation, engagement and integration within the community as well as delivering preventative healthcare and health promotion services.

Sarah will also add this definition to the chat for you to review as needed throughout the session.

The community engaged placement you supervised was conducted over the past two years, and part or all of the placement or placements may have occurred in AccessCampus UL, in local schools or in various services based in the locality. Students will have worked with different community groups and the placement may have involved working with students from other allied health professions and other professionals. Please take a minute or two to recall that experience.

## Focus Group Information Community Liaison Staff

### The topic

Over the past number of years, you will have supervised students from UL as they completed a community engaged placement (CEP) as part of your community service. This study is investigating the learning opportunities experienced during this placement in comparison with other clinical education experiences.

### Ground Rules for the Group

There are some ground rules for our group, but not too many. Firstly, <u>everyone’s experience</u> <u>is valid and valued</u>. We ask that all participants respect different perspectives and viewpoints and contribute to an open sharing of ideas. We would invite you all to share the full extent of your experiences including positives, negatives and areas to work on in the future as this research will inform future practice.

Secondly <u>this focus group is confidential</u> and whilst the session will be recorded participants will be deidentified and anonymised when the results are presented. We ask that all information shared is done so in full confidentiality and remains within the room.

This focus group is about having a conversation with your peers about your experiences. There are some guiding questions and prompts throughout the session but for the most part we will be guided by you and your discussions. Every now and again myself or the moderator (Sarah) will interrupt to ask a question or clarify a point.

## Operational Definitions

For this focus group, community engaged placements refer to placements where students work collaboratively with clients within their local communities. Placement settings are diverse and may include local schools, community facilities or volunteer organisations. When completing a community engaged placement students work with community members in areas to promote and enhance participation, engagement and integration within the community as well as delivering preventative healthcare and health promotion services.

Sarah will also add this definition to the chat for you to review as needed throughout the session.

The community engaged placement you supervised was conducted over the past two years, and part or all of the placement or placements may have occurred in AccessCampus UL, in local schools or in various services based in the locality including your service. Students will have worked with different community groups and the placement may have involved working with students from other allied health professions and other professionals. Please take a minute or two to recall that experience.

## Topic Guide for Student focus group

### Introductions

Before we start, I would invite each of you to Introduce yourself and share something about your background, practice, or general interests.

**Below are the questions for the group,**

1. Can you tell me about your community engaged placement and how you found that experience?

a. <u>Prompt</u> – Was there anything that stood out for you and why?
2. Did you find this placement beneficial, and if so, what are the main benefits that the community engaged placement provided for your learning and practice?

a. <u>Promp</u>t What community groups did you work with and how did you find working with each of them?
b. <u>Prompt</u> What allied health profession students/practitioners did you work with and how did you find working with each of them?
c. <u>Prompt</u> How did you find the supervision model and level of independence on this placement?
d. <u>Prompt</u> If not, why not?
3. What skills and/or competencies did you develop most during the community engaged placement? *(Development of skills or competencies refers to how this placement may have contributed towards developing your proficiency with relation to knowledge or abilities that are necessary to complete your role as an allied health practitioner. Skills/competencies may include communication, teamwork, leadership, clinical reasoning etc)*

a. <u>Prompt:</u> Are there skills/competencies that the community engaged placement added to your practice that traditional clinical placements only would not have given you the opportunity to develop? *(Traditional placements refer to placements that would primarily take place in acute hospital or inpatient settings where patients attend the service with community engaged placements referring to placements where students work collaboratively with clients within their local communities.)*
b. <u>Prompt</u> If yes, what aspect of the placement facilitated this skill/competency development?
c. <u>Prompt</u> Have you kept these skills/competencies up/used it them in other settings?
4. What aspects of the community engaged placement contributed to your learning?

a. <u>Prompt</u> E.g. a chance to put theory into practice, real world/experiential learning, preventative healthcare/health promotion, level of autonomy, working with other AHPs etc.
5. What challenges did the community engaged placement pose?

a. <u>Prompt:</u> Challenges for you?
b. <u>Prompt:</u> Challenges for the community?
6. Have you any suggestions on aspects/considerations for future community engaged placements?

## Topic Guide for the practice education focus group

### Introductions

Before we start, I would invite each of you to Introduce yourself and share something about your background, practice or general interests.

**Below are the questions for the group,**

1. Can you tell me about the community engaged placement(s) you helped to facilitate and how you found that experience?

a. <u>Prompt</u> – Was there anything that stood out for you and why?
2. What were the main benefits and opportunities of supervising students during this placement(s)?

a. <u>Prompt</u> Benefits & opportunities for you as an educator? E.g., supervision model, engagement with community groups, collaboration with other AHPs
b. <u>Prompt</u> Benefits & opportunities for students. E.g., interdisciplinary work, interacting with community groups, autonomy.
c. <u>Prompt</u> Benefits & opportunities for the community?
3. What skills/competencies did you observe students develop most during the community engaged placement? *(Development of skills or competencies refers to how this placement may have contributed towards developing students’ proficiency with relation to knowledge or abilities that are necessary to complete the role as an allied health practitioner. Skills/competencies may include communication, teamwork, leadership, clinical reasoning etc)*

a. <u>Prompt:</u> Are there skills/competencies that the community engaged placement added to student’s practice that traditional clinical placements only would not have given them the opportunity to develop? *(Traditional placements refer to placements that would primarily take place in acute hospital or inpatient settings where patients attend the service with community engaged placements referring to placements where students work collaboratively with clients within their local communities.)*
b. <u>Prompt</u> If yes, what aspect of the placement facilitated this skill/competency development?
c. <u>Prompt</u> Did you notice any unintended education or practice opportunities arise from community partners that contributed towards student learning?
d. <u>Prompt</u> Do you think these skills/competencies are transferable to other settings?
4. What challenges did the community engaged placement pose?

a. <u>Prompt</u> Challenges for you?
b. <u>Prompt</u> Challenges for students?
c. <u>Prompt</u> Challenges for the community?
5. Did you observe any impact the placement had on the community members that the students worked with?

a. <u>Prompt</u> – either short term (during placement) or long-term (after placement finished) impact?
6. Have you any suggestions on aspects/considerations for future community engaged placements?

## Questions for the community liaison staff focus group

### Introductions

Before we start, I would invite each of you to Introduce yourself and share something about your background, practice, or general interests.

**Below are the questions for the group,**

1. Can you tell me about the community the engaged placement you helped to facilitate and how you found that experience?

a. <u>Prompt</u> – Was there anything that stood out for you and why?
2. Did the placement have an impact on your community members/service?

a. <u>Prompt</u> – was there a short-term impact? (Whilst the students were on placement)
b. <u>Prompt</u> – Has there been a longer-term impact? (After students finished placement)
3. How did the students develop during this placement?

a. <u>Prompt:</u> Were there skills/competencies that the community engaged placement added to student’s practice?
b. <u>Prompt</u> Was there differences between students of different levels?
c. <u>Prompt</u> Did you notice specific learning opportunities or gaps that could be addressed? *(Development of skills or competencies refers to how this placement may have contributed towards developing students’ proficiency with relation to knowledge or abilities that are necessary to complete the role as an allied health practitioner. Skills/competencies may include communication, teamwork, leadership, understanding their roles or scope of practice clinical reasoning etc)*
4. What challenges did the community engaged placement pose?

a. <u>Prompt</u> Challenges for you and your community?
b. <u>Prompt</u> Challenges for students?
5. Have you any suggestions on aspects/considerations for future placements in this service or similar services?

## Consolidated criteria for reporting qualitative research (COREQ)

This document consolidates all 32 items of the Consolidated Criteria for Reporting Qualitative Studies and aligns them with evidence from the manuscript.

It includes the mapping table as well as the supplementary text insertions that ensure alignment with COREQ domains: Research Team and Reflexivity, Study Design, and Analysis and Findings.

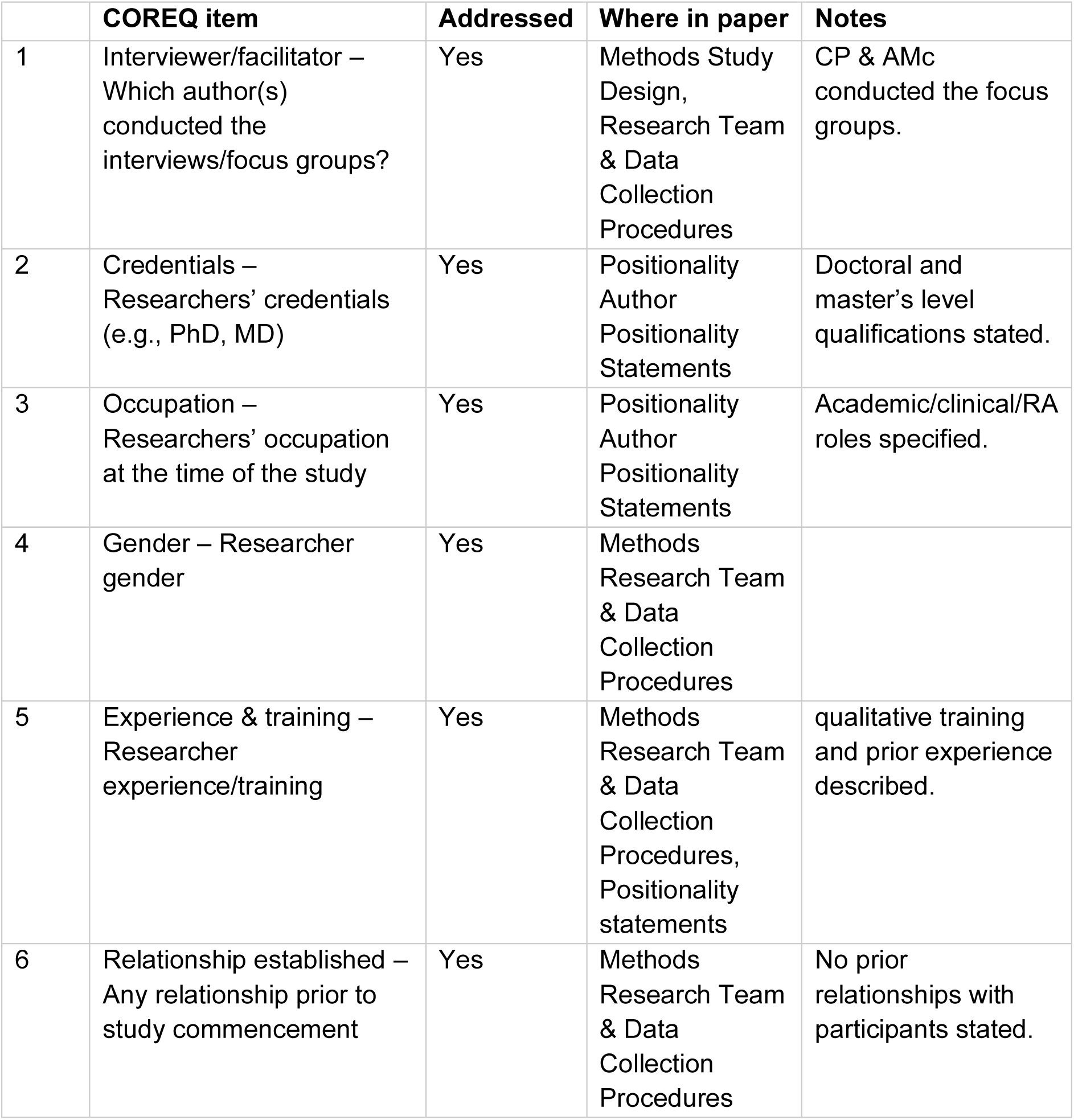
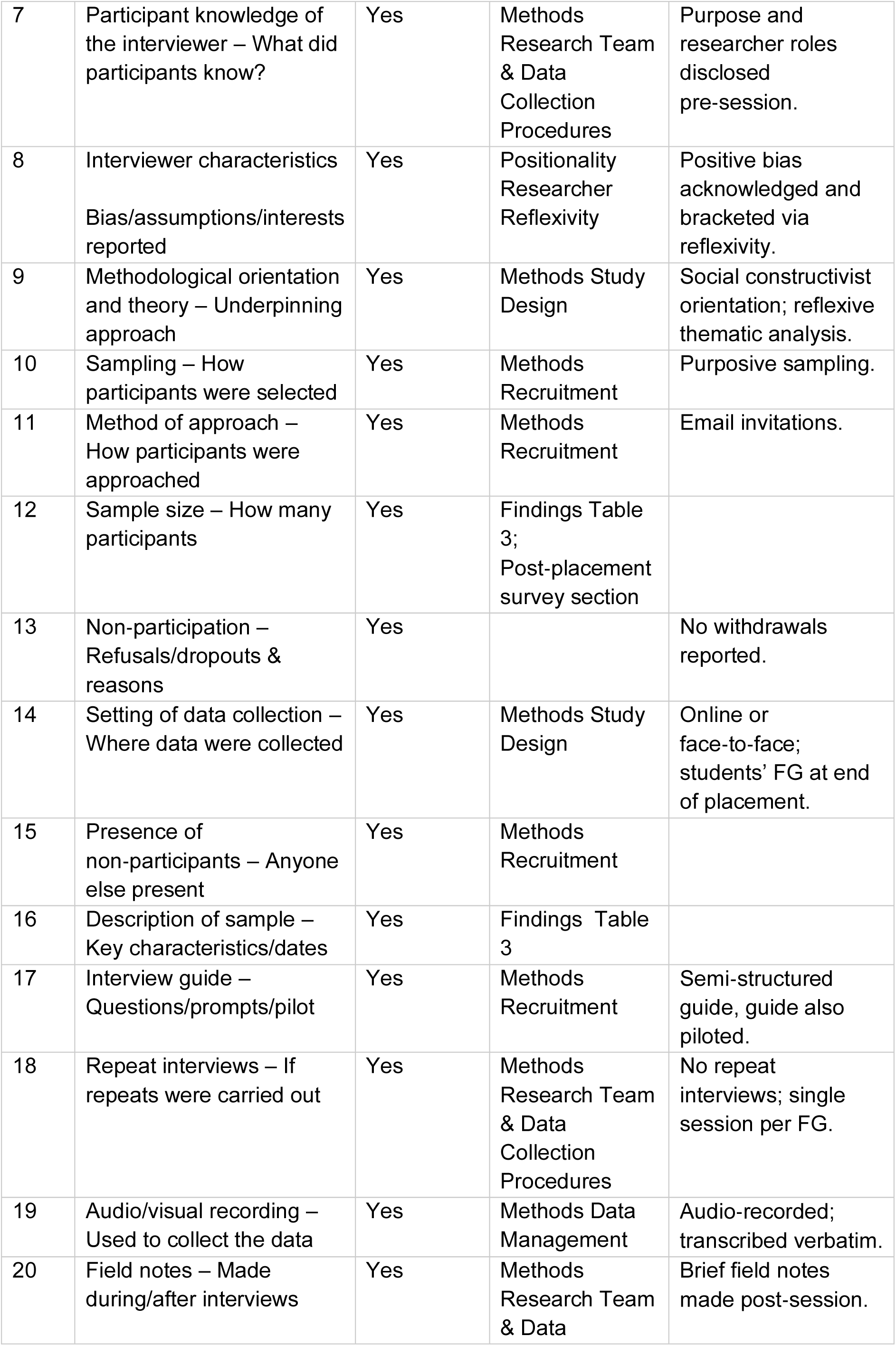
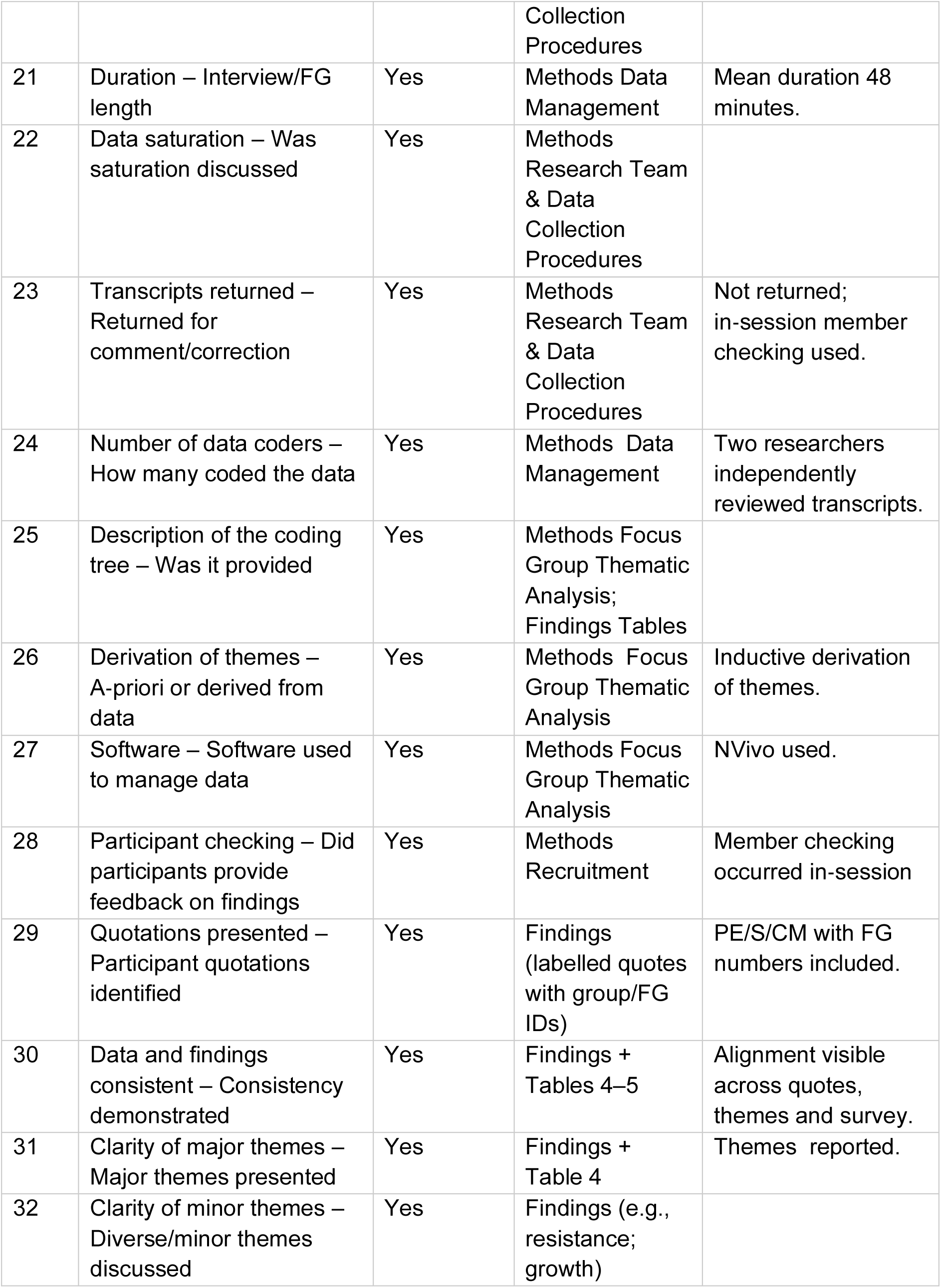

## Positionality and Trustworthiness

The research team brought diverse experience across allied health professional education, community service, social justice, and advocacy. Many had worked as lecturers and had also worked clinically, supervised community-engaged placements, and collaborated closely with community members, while others contributed theoretical expertise in allied health pedagogy. Positionality was acknowledged as pivotal to interpreting data. Reflexivity was maintained throughout via meetings, iterative review, and supervision. Trustworthiness adhered to Lincoln and Guba’s (1985) criteria for credibility, transferability, dependability, and confirmability.

## Researcher Reflexivity

The researchers recognised that their shared commitment to inclusive, community-based practice could introduce positive bias. Reflexive journalling and team dialogue were used to acknowledge and bracket assumptions regarding the benefits of community-engaged placements. As educators who design and assess placements, the team’s positionality was explicitly considered during analysis to reduce the potential for interpretive bias. Reflexivity was further enhanced through cross-checking of coding decisions and independent supervisory oversight to promote transparency and confirmability.

## Supporting information

COREQ

Focus Group guide

## Data Availability

All data produced in the present work are contained in the manuscript.

## Authorship Contributions

- **CP** originated the concept for the study and served as lead author.
- **SD, DOL, TMc, PB, and MOD** collaborated with CP to refine the initial study design.
- **CP and AMc** developed the interview guide and facilitated the focus groups.
- **CP and AMc** completed initial data screening and drafted the preliminary manuscript, with contributions from all team members.
- **ER** undertook the final thematic analysis, refinement of sub-themes, and major revisions. ER served as primary author for manuscript review, synthesis, and submission preparation.
- **CP** – PhD.Physiotherapist and lecturer in SAH, with a concurrent clinical role supporting youth athletes from lower-socioeconomic communities. Seven years’ experience as a community placement tutor. Philosophically grounded in pragmatism and constructivism, drawing on interpretive and critical paradigms. Experienced qualitative researcher and focus group facilitator.
- **SD** – PhD. Physiotherapist and academic staff member in the School of Allied Health (SAH); former CEP participant. Aligns with a constructivist epistemology, viewing meaning as socially co-constructed. Contributed to manuscript review.
- **AMc** – Developmental Officer with a background in sport psychology and social justice. Experienced in participatory practice and community engagement through dance and movement initiatives. Contributed to introductory writing and manuscript review.

**ER** – Occupational Therapist with expertise in qualitative methodology MSc and practice-education models. Conducted the final thematic analysis, refined sub-themes, and finalised the manuscript. Principal author responsible for synthesis, review, and submission.

- **MOD** – Speech and Language Therapist (PhD in Allied Health), Lecturer and Course Director. Engages in community-based participatory research and impact initiatives. Holds a positive orientation toward CEPs and extensive experience in qualitative methods, including focus groups. Contributed to manuscript review.
- **PB** – Occupational Therapist (PhD) with expertise in qualitative methodology, lecturing, and practice-education models. Contributed to review and ensured methodological rigour.
- **DOL** – Lecturer with experience in Allied Health education. Contributed to study design refinement and critical review of the manuscript.
- **TMc** – Occupational Therapist with extensive experience in practice-education and placement coordination. Contributed to manuscript review.
- **Ago** – Occupational Therapist with experience in placement provision and community-engaged practice. Contributed to manuscript review.
- **AGr** – Lecturer and researcher with expertise in qualitative inquiry and Allied Health education. Contributed to manuscript review.
- **BS** –PhD student. Contributed to manuscript review. interest in social justice and community engagement.

