## Supplementary material for "Experiences of a community-engaged placement model: student, educator, and community stakeholder perspectives": COREQ

**Consolidated criteria for reporting qualitative research (COREQ): a 32-item checklist**

This document consolidates all 32 items of the Consolidated Criteria for Reporting Qualitative Studies and aligns them with evidence from the manuscript.

It includes the mapping table as well as the supplementary text insertions that ensure alignment with COREQ domains: Research Team and Reflexivity, Study Design, and Analysis and Findings.

|  | **COREQ item** | **Addressed** | **Where in paper** | **Notes** |
| --- | --- | --- | --- | --- |
| 1 | Interviewer/facilitator – Which author(s) conducted the interviews/focus groups? | Yes | Methods Study Design, Research Team & Data Collection Procedures | CP & AMc conducted the focus groups. |
| 2 | Credentials – Researchers’ credentials (e.g., PhD, MD) | Yes | Positionality Author Positionality Statements | Doctoral and master’s level qualifications stated. |
| 3 | Occupation – Researchers’ occupation at the time of the study | Yes | Positionality Author Positionality Statements | Academic/clinical/RA roles specified. |
| 4 | Gender – Researcher gender | Yes | Methods Research Team & Data Collection Procedures |  |
| 5 | Experience & training – Researcher experience/training | Yes | Methods Research Team & Data Collection Procedures, Positionality statements | qualitative training and prior experience described. |
| 6 | Relationship established – Any relationship prior to study commencement | Yes | Methods Research Team & Data Collection Procedures | No prior relationships with participants stated. |
| 7 | Participant knowledge of the interviewer – What did participants know? | Yes | Methods Research Team & Data Collection Procedures | Purpose and researcher roles disclosed pre‑session. |
| 8 | Interviewer characteristics  Bias/assumptions/interests reported | Yes | Positionality Researcher Reflexivity | Positive bias acknowledged and bracketed via reflexivity. |
| 9 | Methodological orientation and theory – Underpinning approach | Yes | Methods Study Design | Social constructivist orientation; reflexive thematic analysis. |
| 10 | Sampling – How participants were selected | Yes | Methods Recruitment | Purposive sampling. |
| 11 | Method of approach – How participants were approached | Yes | Methods Recruitment | Email invitations. |
| 12 | Sample size – How many participants | Yes | Findings Table 3; Post‑placement survey section |  |
| 13 | Non‑participation – Refusals/dropouts & reasons | Yes |  | No withdrawals reported. |
| 14 | Setting of data collection – Where data were collected | Yes | Methods Study Design | Online or face‑to‑face; students’ FG at end of placement. |
| 15 | Presence of non‑participants – Anyone else present | Yes | Methods Recruitment |  |
| 16 | Description of sample – Key characteristics/dates | Yes | Findings Table 3 |  |
| 17 | Interview guide – Questions/prompts/pilot | Yes | Methods Recruitment | Semi‑structured guide, guide also piloted. |
| 18 | Repeat interviews – If repeats were carried out | Yes | Methods Research Team & Data Collection Procedures | No repeat interviews; single session per FG. |
| 19 | Audio/visual recording – Used to collect the data | Yes | Methods Data Management | Audio‑recorded; transcribed verbatim. |
| 20 | Field notes – Made during/after interviews | Yes | Methods Research Team & Data Collection Procedures | Brief field notes made post‑session. |
| 21 | Duration – Interview/FG length | Yes | Methods Data Management | Mean duration 48 minutes. |
| 22 | Data saturation – Was saturation discussed | Yes | Methods Research Team & Data Collection Procedures |  |
| 23 | Transcripts returned – Returned for comment/correction | Yes | Methods Research Team & Data Collection Procedures | Not returned; in‑session member checking used. |
| 24 | Number of data coders – How many coded the data | Yes | Methods Data Management | Two researchers independently reviewed transcripts. |
| 25 | Description of the coding tree – Was it provided | Yes | Methods Focus Group Thematic Analysis; Findings Tables |  |
| 26 | Derivation of themes – A‑priori or derived from data | Yes | Methods Focus Group Thematic Analysis | Inductive derivation of themes. |
| 27 | Software – Software used to manage data | Yes | Methods Focus Group Thematic Analysis | NVivo used. |
| 28 | Participant checking – Did participants provide feedback on findings | Yes | Methods Recruitment | Member checking occurred in‑session |
| 29 | Quotations presented – Participant quotations identified | Yes | Findings (labelled quotes with group/FG IDs) | PE/S/CM with FG numbers included. |
| 30 | Data and findings consistent – Consistency demonstrated | Yes | Findings + Tables 4–5 | Alignment visible across quotes, themes and survey. |
| 31 | Clarity of major themes – Major themes presented | Yes | Findings + Table 4 | Themes reported. |
| 32 | Clarity of minor themes – Diverse/minor themes discussed | Yes | Findings (e.g., resistance; growth) |  |

a. Prompt If yes, what aspect of the placement facilitated this skill/competency development?
b. Prompt Did you notice any unintended education or practice opportunities arise from community partners that contributed towards student learning?
c. Prompt Do you think these skills/competencies are transferable to other settings?
